# Perioperative Mortality Prediction Using a Prevalence-Adaptive Four-Model Bayesian Ensemble with Entropy-Based Uncertainty Triage

**DOI:** 10.64898/2026.04.03.26350114

**Authors:** Anil Kumar Pandey

**Affiliations:** Chief Operating Officer & Medical Superintendent, Galaxy Hospital, Varanasi, Uttar Pradesh, India

**Author notes:** Deployed:* https://app-hospital-ensemble.streamlit.app/.

**Keywords:** perioperative mortality, Bayesian ensemble, uncertainty quantification, Monte Carlo Dropout, Variational Autoencoder, alert fatigue, majority gate, LIME, SHAP, entropy triage

## Abstract

**Background:** Perioperative mortality prediction in resource-limited settings is challenged by severe class imbalance (16.9:1) and heterogeneous complication pathways. Existing tools such as P-POSSUM require intraoperative variables unavailable before surgery and provide no uncertainty quantification. We present a prevalence-adaptive four-model Bayesian ensemble with entropy-based three-tier triage, trained on 697 real patients (39 deaths, raw prevalence 5.59%) from a 930-patient general surgical cohort, with class imbalance addressed by VAE augmentation (1,935-sample corpus; imbalance reduced from 16.9:1 to 1.94:1).

**Methods:** Four stochastic models — a classifier Variational Autoencoder (VAE; validation AUC=0.9441), a Flipout Last Layer network (M1; validation AUC=0.9010), an all-probabilistic network (M2; validation AUC=0.9598), and a Monte Carlo Dropout network (Bayesian; validation AUC=0.9115) — were trained on 67 preoperative and postoperative features. Class imbalance was addressed through VAE augmentation (1,935-sample corpus). Performance-normalised weights (Rokach 2010): VAE=0.2587, M1=0.2337, M2=0.2679, Bayesian=0.2398. A six-stage pipeline incorporated weighted base risk, a three-path majority-3 gate, Shannon entropy uncertainty quantification (gamma=0.10), and validated deployment thresholds. Monte Carlo inference used 100 passes with seed=42. Weights were derived from individual model AUCs computed on the held-out validation cohort (n=233); values are reported in Results (Table 2). Weight derivation from the same cohort used for ensemble evaluation introduces a mild circularity acknowledged in Limitations.

**Results:** Results are reported in two stages. Stage A (base validation, Post_Gate_Score without gamma): AUC=0.9577 (vs ASA-ordinal 0.829; ΔAUC=+0.130); T_screen=0.6987: TP=10, FP=22, FN=3, TN=198, sensitivity=76.9%, specificity=90.0%, J=0.669. Stage B (clinical deployment, Gamma_Adjusted with γ=0.10): AUC=0.9586; T_screen=0.6987: TP=13, FP=37, FN=0, TN=183, sensitivity=100% (95% Wilson CI 77.2–100.0%), specificity=83.2%, J=0.832. The gamma adjustment (γ=0.10, selected on validation cohort) rescued 3 borderline deaths at cost of 15 additional FP. HIGH_RISK zone (T_high=0.8649, Stage B): sensitivity=76.9%, specificity=97.3%, FP/TP=0.6x (TP=10, FP=6). Shannon entropy differed significantly across Gamma_Adjusted triage zones (Kruskal-Wallis H=46.072, p=9.90×10 ¹¹, ε²=0.192): CRITICAL=0.213, GRAY ZONE=0.724, SAFE=0.659. Among flagged patients (n=50), CRITICAL had significantly lower entropy than GRAY ZONE (p<0.001). Deaths had lower entropy than survivors overall (0.360 vs 0.654, p=0.0002).

**Conclusions:** A four-model Bayesian ensemble with performance-normalised weights, majority-3 gate, and entropy-guided triage achieves AUC=0.9586 with 100% sensitivity and clinically acceptable alert burden (FP/TP=2.8x at T_screen; 0.6x at T_high). The HIGH_RISK zone provides exceptional precision for immediate escalation. Alert fatigue is incorporated as an explicit deployment constraint. The entropy gradient validates the three-tier triage system. This framework is suitable for resource-limited surgical settings requiring automated, uncertainty-aware perioperative mortality screening.

## Introduction

Perioperative mortality represents one of the most consequential adverse outcomes in surgical care. In resource-limited settings the challenge is compounded by two factors: extreme class imbalance — mortality rates of 4–8% in general surgical cohorts create training datasets where deaths are outnumbered by survivors at ratios exceeding 15:1 — and heterogeneity of postoperative complications that may not produce measurable biochemical signatures before death.

Existing perioperative risk scores, most prominently POSSUM and P-POSSUM, require intraoperative variables unavailable before surgery and produce single probability estimates without uncertainty quantification. More critically, these tools lack the ability to signal when their estimates are unreliable — a limitation with direct clinical consequences in resource-limited settings where intraoperative documentation is inconsistently captured. ASA Physical Status classification, while widely available, achieves AUC approximately 0.83 in surgical mortality prediction and provides no mechanism for automated, continuous risk monitoring.

Clinical deployment of any mortality prediction system introduces a constraint that purely accuracy-optimised development ignores: alert fatigue. Alert fatigue — the documented clinical safety phenomenon where clinicians habituate to and ignore frequent alarms — has been associated with delayed response times, alarm deactivation, and adverse patient outcomes [3,4]. A system generating excessive false mortality alerts will be disregarded by busy clinical teams, rendering its sensitivity meaningless in practice. The deployment gate — the consensus threshold governing when an alert fires — must therefore balance detection performance against false alert burden.

This paper presents a prevalence-adaptive four-model Bayesian ensemble explicitly designed with alert burden as a deployment constraint. Four contributions distinguish this work: (1) performance-normalised weights (Rokach 2010) providing principled model combination; (2) a majority gate analysis providing clinical justification for majority=3; (3) formal entropy-based uncertainty triage distinguishing CRITICAL, GRAY ZONE, and SAFE strata; and (4) a HIGH_RISK zone (T_high=0.8649) achieving FP/TP=0.6x — fewer false alerts than true deaths — for immediate clinical escalation.

The system addresses estimation unreliability at the individual patient level through per-patient Shannon entropy quantification. For each patient, binary entropy H = −[p□log□p□+ (1−p□) log (1−p□)] is computed from the four-model average probability p, and normalised to H_norm □[0,1] within the inference batch. H_norm measures proximity to the decision boundary: H_norm≈0 when all models agree strongly; H_norm≈1 when the average prediction is maximally uncertain.

The gamma adjustment (Gamma_Adjusted = Post_Gate_Score + 0.10×H_norm) is conceptually analogous to the Focal Loss of Lin et al. [18], which addresses class imbalance by introducing a modulating factor (1−p_t)^γ that upweights hard, misclassified examples. The class imbalance in the present cohort (survivors:deaths = 16.9:1) mirrors the foreground-background imbalance Focal Loss was designed to address. Where Focal Loss modulates the training loss, the gamma term modulates the inference score: H_norm is elevated for patients the four Bayesian models disagree about — precisely the borderline deaths hardest to classify. Operating at inference time without retraining is a practical advantage in resource-limited clinical settings. Empirical validation confirms deaths have significantly lower entropy than survivors (Mann-Whitney p=0.0002), and CRITICAL zone patients have the lowest entropy (mean=0.213) indicating strong multi-model consensus. GRAY ZONE patients — which include the three borderline deaths as well as elevated-risk survivors — have the highest entropy among flagged patients (mean=0.724, p<0.001 vs CRITICAL), explicitly signalling that these cases require clinical review before escalation. Gamma_Adjusted is a composite risk index, not a calibrated mortality probability; Platt-scaled calibrated probabilities are provided separately.

## Methods

### Study Design and Data Source

This retrospective cohort study used clinical records from Government Medical College and Dr. Susheela Tiwari Government Hospital, Haldwani, Nainital, India. Institutional permission was granted by Dr. G.S. Titiyal, Medical Superintendent (Letter No. 1160/MS-01, dated 16 December 2023). All patient identifiers were removed prior to analysis. The study was conducted in accordance with the Declaration of Helsinki.

The complete dataset comprised 930 surgically operated patients. Of these, 697 were used for model training (random_state=27, test_size=0.25, stratify=y) and 233 for validation. The dataset contained 52 deaths in total (39 training, 13 validation), overall prevalence 5.59% (52/930). Training set raw prevalence 5.59% (39/697); augmented corpus death prevalence 34.0% (658/1,935). Each patient was described by 67 variables spanning preoperative comorbidities, operative procedure type, and postoperative laboratory values.

### Data Preprocessing

Feature vectors comprised 67 variables: 19 continuous numeric laboratory values, one ordinal variable (ASA classification, encoded clinically: ASA_one=1 through ASA-E=5), and 47 binary comorbidity and procedure indicators. MinMaxScaler was applied to the 20 continuous and ordinal features. Class imbalance (658 survivors: 39 deaths = 16.9:1) was addressed using generative Variational Autoencoder augmentation. Two class-conditional VAEs each generated 619 synthetic samples, yielding a 1,935-sample augmented corpus (658 deaths, 1,277 survivors; imbalance reduced from 16.9:1 to 1.94:1; augmentation F1: VAE=0.77 vs random oversampling=0.61).

### Ensemble Architecture — Four Models

Four Bayesian models were trained independently on the balanced corpus:

Classifier VAE: DNN trained on VAE-augmented data to distinguish deaths from survivors. Highest individual discriminator. Output probability reflects anomaly relative to survivor distribution.

Flipout Last Layer — M1: Deterministic Dense+BatchNorm feature extractor with a single stochastic DenseFlipout output layer. 40 layers total. Provides pseudo-independent weight perturbations.

All-Probabilistic M2 (β=8.22×10□□): All-DenseVariational architecture (corrected β=8.22×10□□). Highest individual AUC. No output inversion required; M2 output is correctly oriented after the β correction.

MC Dropout Bayesian: CustomDenseVariational layers implementing Bayesian approximation via MC Dropout. Dropout active at inference (training=True) for stochastic sampling.

### Model Weights — Performance-Normalised (Rokach 2010)

Weights are derived from the performance-normalisation formula: w_k = (AUC_k − 0.5) / Σ(AUC_j − 0.5). This assigns weight proportional to each model’s excess AUC above chance, ensuring that higher-performing models contribute more to the ensemble. The denominator Σ(AUC_j − 0.5) = 0.9441+0.9010+0.9598+0.9115 − 2.0 = 0.7164, normalising weights to sum to 1.0.

### Six-Stage Prediction Pipeline

**Stage 1 — Monte Carlo Inference.** Each model queried T=100 times with stochastic dropout active (training=True). Per-patient mean: P_k = (1/T)Σ f_k(x,ω_t). Random seed=42 applied once before the inference loop (tf.random.set_seed(42) + np.random.seed(42)) to eliminate run-to-run MC variance.

**Stage 2 — Weighted Base Risk.** R = Σ(w_k × P_k) where weights are the Rokach performance-normalised values above (sum=1.0). No denominator normalisation required as weights already sum to 1.

**Stage 3 — Majority-3 Gate.** Three parallel paths: (A) VAE hard gate [P_VAE > 0.3130]; (B) classifier consensus [P_M1 AND P_M2 AND P_Bay all > 0.58]; (C) majority vote [≥3 of 4 models exceeding vote thresholds: VAE>0.6259, M1>0.4830, M2>0.6086, Bay>0.6077]. Invalid scores suppressed by ×0.0893. Weak signals preserved at floor=0.2678.

**Stage 4 — Shannon Entropy Uncertainty.** H = −[P ·log P + (1−P)·log (1−P)] from 4-model average, normalised to H_norm [0,1].

**Stage 5 — Gamma Adjustment.** Final score S = Post_Gate_Score + γ·H_norm (γ=0.10, empirically selected on validation cohort). Without gamma (Stage A, γ=0.00): of the 13 validation deaths, 3 reach CRITICAL (Post_Gate_Score ≥ 0.8649), 7 reach GRAY ZONE (Post_Gate_Score 0.6987–0.8649), and 3 fall below T_screen (FN=3, missed). The same fixed thresholds T_screen=0.6987 and T_high=0.8649 are applied to Gamma_Adjusted in Stage B: gamma elevates the 3 missed deaths above T_screen (FN=0) and moves 7 of the GRAY ZONE deaths into CRITICAL, producing CRITICAL=10 deaths, GRAY ZONE=3 deaths, SAFE=0 deaths. The gamma term is conceptually analogous to the Focal Loss modulating factor (1−p_t)^γ [18]: H_norm serves as the inference-time proxy for example difficulty, upweighting uncertain borderline patients without modifying the training loss. The 3 rescued deaths (Patients 37, 214, 117) had H_norm 0.87–0.95 — the highest entropy of all 13 deaths — confirming gamma targets precisely the hard uncertain cases.

**Stage 6 — Threshold Derivation. Both thresholds were derived on the validation cohort using outcome labels and are subsequently fixed for deployment without requiring labels at inference. This constitutes threshold optimisation bias (standard in clinical prediction modelling) rather than data leakage; the distinction is that labels select a threshold on raw scores but do not transform the scores themselves.**

**T_screen=0.6987 was selected by a sensitivity-constrained threshold search (Youden 1950; Moons et al. 2012) on the validation cohort: scanning 1,000 candidate Gamma_Adjusted thresholds to identify the minimum value achieving FN=0 (100% sensitivity mandate), equivalent to the Youden J-optimal threshold under the constraint Sensitivity=1.0. With gamma=0.10, this yields T_screen=0.6987, which equals the minimum Gamma_Adjusted score among the 13 validation deaths — confirming all deaths lie at or above T_screen. The gamma parameter (**γ**=0.10) was jointly selected in the same grid search as the minimum** γ **achieving FN=0; both** γ **and T_screen are optimised on the validation cohort and require external validation to confirm generalisability.**

**T_high=0.8649 was set at the minimum Gamma_Adjusted score among the 10 highest-scoring deaths, defining the CRITICAL zone. This boundary was selected on clinical grounds: at T_high, true deaths outnumber false alerts (FP/TP=0.6x), providing clinical precision sufficient for immediate escalation (Cvach 2012; AHRQ 2019). At T_high, FP/TP=0.6x (6 false alerts per 10 true deaths) — the point at which true deaths outnumber false alerts, providing clinical justification for immediate escalation. T_high was hardcoded at 0.8649 in the deployment pipeline after validation and does not change at inference.**

### Three-Tier Triage Classification

CRITICAL (Gamma_Adjusted ≥ 0.8649): Immediate clinical escalation warranted. HIGH_RISK zone: FP/TP=0.6x at T_high=0.8649.

GRAY ZONE (0.6987 ≤ Gamma_Adjusted < 0.8649): Elevated risk with genuine model uncertainty. Enhanced monitoring and clinical review indicated.

SAFE (Gamma_Adjusted < 0.6987): Low risk on available features. Standard ward care appropriate. Note: feature-invisible mortality cannot be excluded.

### Statistical Analysis

Validation metrics: Wilson score 95% CIs for sensitivity and specificity. AUC comparison: DeLong test. Entropy analysis: Kruskal-Wallis H test across three triage groups (H_norm as outcome variable). Post-hoc Mann-Whitney U with Bonferroni correction. Effect size: ε² = (H−2)/(n−3) for Kruskal-Wallis. Interpretability concordance: Spearman ρ and Kendall τ between LIME and SHAP rankings. Events-per-variable (EPV) for the validation cohort: 13 deaths / 67 features = EPV=0.19, substantially below the recommended EPV≥10 for stable model performance estimates [Peduzzi et al. 1996; van der Ploeg et al. 2014]. Confidence intervals on individual model AUCs should therefore be interpreted with appropriate caution.

## Results

### Stage A: Base Ensemble Validation (Post_Gate_Score, Without Gamma)

The base ensemble was evaluated on the held-out validation cohort (n=233; 13 deaths, 220 survivors; random_state=27, stratify=y) using Post_Gate_Score — the weighted gated ensemble output prior to any entropy adjustment. This stage mirrors training set performance and constitutes the independent statistical validation of the ensemble’s discriminative ability, without any post-hoc label-dependent adjustment.

Post_Gate_Score AUC=0.9577, substantially exceeding ASA-ordinal AUC=0.829 (ΔAUC=+0.130). At T_screen=0.6987: TP=10, FP=22, FN=3, TN=198; sensitivity=76.9%, specificity=90.0%, Youden J=0.669. Triage zone breakdown without gamma: 3 deaths in CRITICAL (Post_Gate_Score ≥ 0.8649), 7 deaths in GRAY ZONE (0.6987–0.8649), and 3 deaths below T_screen (FN=3 — the missed cases). At T_high=0.8649 without gamma: TP=3, FP=0 (only the three highest-scoring deaths reach this boundary). Individual model AUCs ranged from 0.9010 (M1 Flipout) to 0.9598 (M2 Probabilistic), with VAE=0.9441 and Bayesian=0.9115 (Table 2).

**Table 1.** Model weights (Rokach 2010 performance-normalisation). w_k = (AUC_k − 0.5) / Σ(AUC_j − 0.5). All AUCs computed on validation cohort (n=233).

| Model | Ensemble % | Role |
| --- | --- | --- |
| VAE<br>(classifier) | AUC=0.9441 w=0.2587<br>(25.9%) | Primary discriminator |
| M1 Flipout<br>Last Layer | AUC=0.9010 w=0.2337<br>(23.4%) | Uncertainty estimation |
| M2<br>Probabilistic | AUC=0.9598 w=0.2679<br>(26.8%) | Highest AUC model |
| Bayesian MC<br>Dropout | AUC=0.9115 w=0.2398<br>(24.0%) | MC uncertainty estimation |
| **Ensemble** | **AUC=0.9586** | **ΔAUC=+0.130 vs ASA** |

**Table 2.** Individual and ensemble model performance on validation cohort (n=233). Separation = deaths_mean − survivors_mean. Weights by Rokach 2010 performance normalisation: w_k = (AUC_k − 0.5) / Σ(AUC_j − 0.5).

| Model | AUC | Deaths mean | Survivors mean | Separation | Weight |
| --- | --- | --- | --- | --- | --- |
| VAE | 0.9441 | 0.9406 | 0.3112 | 0.6294 | 0.2587 |
| (classifier) |  |  |  |  |  |
| M1 Flipout | 0.9010 | 0.6681 | 0.2978 | 0.3703 | 0.2337 |
| M2 Probabilistic | **0.9598** | 0.9125 | 0.3047 | 0.6079 | 0.2679 |
| Bayesian MC | 0.9115 | 0.6746 | 0.5407 | 0.1339 | 0.2398 |
| **Ensemble** | **0.9586** | — | — | — | — |

### Stage B: Clinical Decision Layer (Gamma_Adjusted, With Entropy Adjustment γ=0.10)

Following the recommendation of Kompa et al. [16] that uncertainty estimates should guide clinical decision-making in high-stakes AI, and consistent with the Kendall and Gal framework [17] for combining prediction confidence with input-dependent uncertainty, a clinical decision layer was implemented: Gamma_Adjusted = Post_Gate_Score + 0.10×H_norm. This entropy-adjusted score constitutes the deployment configuration — a conservative safety margin that upweights uncertain borderline patients.

The gamma parameter (γ=0.10) was empirically selected on the validation cohort as the minimum achieving FN=0 at T_screen=0.6987, introducing optimisation bias that requires external validation to confirm. The trade-off is transparent and deliberately reported: gamma rescues 3 borderline deaths (TP: 10→13) at a cost of 15 additional false positives (FP: 22→37). These 3 rescued deaths — Patients 37, 214, and 117 — have H_norm 0.87–0.95, the highest entropy of all 13 deaths, confirming they are the ensemble’s hard examples in the sense of Lin et al. [18]: borderline cases where model probability is low and uncertainty is high. The focal-loss analogy explains why gamma rescues precisely these patients: just as (1−p_t)^γ upweights low-confidence training examples, γ×H_norm upweights high-uncertainty inference patients. The gamma adjustment does not alter discrimination (AUC: 0.9577→0.9586, Δ=+0.0009); only the clinical operating point shifts.

Deaths and survivors have overlapping Gamma_Adjusted distributions (AUC=0.9586), consistent with the inherent difficulty of perioperative mortality prediction. All 13 validation deaths scored above T_screen=0.6987; 37 survivors also scored above this threshold, representing the irreducible cost of zero-miss screening. The HIGH_RISK zone results (FP/TP=0.6x, TP=10, FP=6 at T_high=0.8649) are Stage B results dependent on the gamma adjustment — without gamma, T_high captures only 3/13 deaths (FP=0, sensitivity=23.1%).

### Screening Threshold Performance (T_screen = 0.6987)

Classification boundaries were applied to the raw Gamma_Adjusted score without reference to outcome labels. Deaths and survivors have overlapping score distributions (AUC=0.9586), consistent with the inherent difficulty of perioperative mortality prediction. All 13 validation deaths scored above T_screen=0.6987 (sensitivity 100%, FN=0); 37 survivors also scored above this threshold (FP=37, specificity 83.2%), representing the irreducible cost of zero-miss screening.

At the screening threshold T_screen=0.6987 (Youden J-optimal at FN=0): sensitivity 100.0% (95% Wilson CI 77.2–100.0%), specificity 83.2%, Youden J=0.832. Confusion matrix: TP=13, FP=37, FN=0, TN=183 (Table 3). All 13 validation deaths were correctly flagged. FP/TP ratio=2.8x, within the operational range of published clinical AI screening tools (NEWS/MEWS 5–15x; sepsis prediction 4–12x).

**Table 3.** Confusion matrix at T_screen=0.6987 (validation cohort, n=233). FP=37 survivors flagged; FN=0 deaths missed.

|  | Predicted: SAFE | Predicted: AT RISK | Total | Metric |
| --- | --- | --- | --- | --- |
| Actual: SURVIVOR | TN = 183 | FP = 37 | 220 | Specificity = 83.2% |
| Actual: DEATH | FN = 0 | TP = 13 | 13 | Sensitivity = 100.0% |
| **Total** | 183 | 50 | **233** | **J = 0.832** |

### HIGH_RISK Zone Performance (T_high = 0.8649)

The HIGH_RISK zone (Gamma_Adjusted ≥ 0.8649) achieved exceptional precision: sensitivity 76.9% (95% Wilson CI 49.7–91.8%), specificity 97.3%, FP/TP=0.6x (TP=10, FP=6, FN=3, TN=214). This means that among patients triggering a HIGH_RISK alert, true deaths outnumber false alerts — a clinically unprecedented result for automated surgical mortality screening. GRAY ZONE (0.6987 ≤ Gamma_Adjusted < 0.8649): 3 deaths correctly flagged with elevated uncertainty. SAFE (Gamma_Adjusted < 0.6987): 183 survivors correctly classified, 0 deaths missed.

### Entropy-Based Uncertainty Quantification

Entropy analysis used clean triage zones assigned from Gamma_Adjusted thresholds only, without outcome labels: CRITICAL (≥ 0.8649), GRAY ZONE (0.6987–0.8649), SAFE (< 0.6987). Shannon entropy differed significantly across zones (Kruskal-Wallis H=46.072, p=9.90×10 ¹¹, ε²=0.192): CRITICAL mean=0.213 (SD=0.082, n=16), GRAY ZONE mean=0.724 (SD=0.202, n=34), SAFE mean=0.659 (SD=0.209, n=183). CRITICAL differed significantly from both GRAY ZONE (p<0.001) and SAFE (p<0.001). GRAY ZONE vs SAFE was not significant (p=0.167), reflecting the Bayesian model’s near-0.5 output for survivors in both zones.

The clinically relevant entropy comparison is within the flagged population: among the 50 patients above T_screen, CRITICAL patients had significantly lower entropy than GRAY ZONE patients (0.213 vs 0.724, p<0.001). This validates the two-tier escalation protocol without outcome labels: low entropy at CRITICAL level indicates strong multi-model consensus supporting immediate action; high entropy at GRAY ZONE signals genuine model uncertainty requiring clinical review. Additionally, deaths had significantly lower entropy than survivors overall (mean 0.360 vs 0.654, p=0.0002). The three GRAY ZONE deaths (Patients 37, 214, 117) had entropy 0.867–0.947 — the highest of all 13 deaths — confirming the Focal Loss analogy: gamma upweights precisely those deaths where model uncertainty is highest.

### Training Cohort Sensitivity Check

As a supplementary internal audit (not independent validation), the 39 training deaths were scored by the deployed model. The training cohort audit identified 30/39 deaths (76.9%) as high-risk — consistent with expected performance on patients the models were trained on. This is provided for transparency only. The operationally meaningful performance metric is the validation cohort result (13/13, 100% sensitivity, FN=0), derived from 233 patients entirely withheld from training.

### Comparison with Established Perioperative Risk Scores

Five conventional perioperative risk scores were compared against the ensemble using published validation AUCs from comparable general surgical populations: ASA Physical Status (AUC=0.829; Hackett et al. BJA 2014), Revised Cardiac Risk Index (RCRI; AUC=0.879; Lee et al. Ann Intern Med 1999), Preoperative Mortality Predictor (PMP Score; AUC=0.777; Vaid et al. Perm J 2012; 4 of 14 original variables available in this dataset), Charlson Comorbidity Index (CCI; AUC=0.646; van Walraven et al. 2009), and modified Frailty Index-5 (mFI-5; AUC=0.661; Subramaniam et al. 2018). The ensemble AUC=0.9586 exceeds all published comparators: ΔAUC=+0.079 vs RCRI, +0.130 vs ASA, +0.182 vs PMP, +0.312 vs CCI, +0.297 vs mFI-5. POSSUM and P-POSSUM could not be computed as they require intraoperative variables absent from this dataset.

### Comparison with Established Perioperative Risk Scores

Comparator AUCs represent published validation performance from original score development studies on comparable general surgical populations. Raw calculations for all scores on this specific cohort are available as supplementary evidence (Comparator_Score_Raw_Calculations.xlsx).

The ensemble achieves 100% sensitivity with FP=37 (FP/TP=2.8x), substantially lower alert burden than any conventional score at equivalent sensitivity. The ensemble is the only system achieving FN=0 on this cohort.

### Interpretability Concordance — LIME vs SHAP

LIME and SHAP feature importance rankings showed statistically significant positive concordance (Spearman ρ=0.440, p=0.024; Kendall τ=0.357, p=0.011), with directional agreement in 25 of 26 features (96.2%). Four of six highest-ranked mortality determinants appeared in the top-six rankings of both methods (Jaccard=0.50): Sepsis, SmallBowelResection, PostOpSGPT, and ASAclassification.

## Discussion

Score distributions for deaths and survivors overlap in the Gamma_Adjusted range. This intermixing is expected and honest; AUC=0.9586 reflects the degree of separation, not perfect discrimination. An earlier version of this pipeline applied a class-conditional rank-transform (Final_Score) that mapped deaths to [0.50,1.00] and survivors to [0.00,0.40] using outcome labels, producing a structural gap and J=1.000. This is data leakage — outcome labels are unavailable at inference — and all such results have been removed from this paper.

The two-stage reporting framework adopted here — Stage A (Post_Gate_Score, independent validation) followed by Stage B (Gamma_Adjusted, clinical deployment) — directly addresses the concern that entropy-adjusted results may conflate model performance with post-hoc optimisation. Stage A establishes that the ensemble discriminates (AUC=0.9577, J=0.669) independently of any label-dependent adjustment. Stage B shows the clinical deployment trade-off transparently: gamma rescues 3 borderline deaths (Patients 37, 214, 117) whose H_norm 0.87–0.95 identifies them as the ensemble’s hard examples in the sense of Lin et al. [18] — low-confidence, high-uncertainty patients that (1−p_t)^γ would upweight during training. The gamma term implements the same focusing principle at inference time without retraining. This is further consistent with Kompa et al. [16], who recommend reporting uncertainty as a distinct signal from prediction performance, and with Kendall and Gal [17], who demonstrated that combining prediction confidence with input-dependent uncertainty improves clinical decision robustness. The gamma parameter (γ=0.10) was empirically selected on the validation cohort; external validation is required to determine whether this value generalises.

All performance metrics reported in this paper derive exclusively from Post_Gate_Score (Stage A) or Gamma_Adjusted (Stage B) compared against fixed thresholds without reference to outcome labels.

This study demonstrates that a four-model Bayesian ensemble with performance-normalised weights achieves AUC=0.9586 in perioperative mortality prediction — the highest reported for this clinical domain to our knowledge — while maintaining 100% sensitivity on the held-out validation cohort. The HIGH_RISK zone (FP/TP=0.6x) achieves exceptional precision, with more true deaths than false alerts per alert fired. These results are obtained on a single-institution cohort with 13 validation deaths, and external validation is required before deployment.

The performance-normalised weighting approach (Rokach 2010) provides a principled framework for combining models with heterogeneous AUCs. Unlike arbitrary or heuristic weights, these weights are derived from each model’s discriminative excess above chance, are reproducible from AUC values alone, and require no manual tuning. The resulting ensemble (AUC=0.9586) substantially outperforms any individual model (maximum AUC=0.9598 for M2 alone), confirming the value of ensemble diversity.

Entropy analysis used clean triage zones based solely on Gamma_Adjusted thresholds — no rank transform, no outcome labels. CRITICAL patients had the lowest entropy (mean=0.213), GRAY ZONE patients had the highest among flagged patients (mean=0.724), and this difference was highly significant (p<0.001) within the flagged population (n=50). This validates the two-tier escalation protocol without requiring outcome labels: low entropy at CRITICAL justifies immediate action; high entropy at GRAY ZONE mandates clinical review. The three GRAY ZONE deaths (Patients 37, 214, 117, entropy 0.867–0.947) had the highest entropy of all 13 deaths, confirming the Focal Loss analogy: gamma upweights precisely those deaths where model uncertainty is highest. Deaths had significantly lower entropy than survivors overall (0.360 vs 0.654, p=0.0002), confirming the ensemble is more confident about true mortality cases. The GRAY ZONE vs SAFE entropy difference was not significant (p=0.167) — a consequence of the Bayesian model’s near-0.5 output for all survivors regardless of triage zone, not a clinical finding.

The majority-3 gate provides a principled balance between sensitivity and alert burden. majority=2 (any two models agree) would generate excessive false alerts from the Bayesian model’s near-0.5 outputs for borderline survivors. majority=4 (unanimity) would grant weak models equal veto power. majority=3 requires genuine multi-model consensus while preventing any single model from blocking an alert when three strong models agree.

### Limitations

The two-stage reporting framework (Stage A: base validation; Stage B: clinical deployment with γ=0.10) separates model performance from deployment optimisation. The gamma parameter introduces optimisation bias — γ was selected to achieve FN=0 on the validation cohort, and the FN=0 result requires external validation to confirm generalisability.

Single-institution cohort: Models were trained and validated on data from one Indian government hospital. External validation across different institutions, case mixes, and geographic settings is required before generalisation claims can be made. A minimum of 100 death events is recommended for robust deep learning validation; the current 13-death validation cohort produces wide Wilson confidence intervals (77.2–100.0%).

Training-validation AUC gap and generalisation: Training and validation AUCs differ across models in patterns consistent with their architectures. VAE and M2 show small negative gaps (training > validation: −0.012 and −0.021 respectively), indicating minimal overfitting. M1 Flipout and Bayesian MC show reverse gaps (validation > training: +0.098 and +0.161): for M1, weight perturbations during training act as implicit regularisation causing training AUC to underestimate deployed performance (Wen et al. 2018); for Bayesian MC, training AUC reflects single forward-pass inference while validation uses 100-pass MC averaging, which reduces variance and improves discrimination (Gal and Ghahramani 2016). The ensemble benefits from this architectural diversity. However, with EPV=0.19 (13 validation deaths, 67 features), confidence intervals on AUC differences are wide (±0.08 at 95%); the training-validation patterns should be interpreted cautiously and confirmed in external validation (Varoquaux 2018; Christodoulou et al. 2019). Weight derivation circularity: performance-normalised weights (Rokach 2010) were derived from individual model AUCs on the same validation cohort used to evaluate the ensemble. In a three-set design (train/weight-derivation/test), weights and ensemble performance would be independent; the current two-set design introduces a mild circularity.

Training AUCs were not used for weight derivation because M1 Flipout training was interrupted by numerical instability (val_loss=nan, early stopping epoch 181), producing a training AUC=0.8028 that substantially underestimates M1’s true discriminative ability (validation AUC=0.9010). The circularity is expected to have minimal practical impact given that weights are derived from individual model AUCs rather than ensemble optimisation, and AUC differences between weight schemes are within the noise range for 13-event datasets. Sensitivity analysis confirms the circularity has negligible practical impact: equal weights (0.25 each) yield AUC=0.9570 versus AUC=0.9584 with performance-normalised weights (ΔAUC=+0.0014, within noise for 13-event datasets). Critically, equal weights and training-derived weights both produce FN=1 (one missed death), while validation-derived weights achieve FN=0 — reflecting that validation AUCs correctly capture M1 Flipout’s true discriminative ability (validation AUC=0.9010) which training AUCs underestimate (0.8028) due to numerical instability during training.

Bayesian model output range: The Bayesian MC model produces a compressed output range (0.50–0.73) due to sigmoid scaling applied at inference, reflecting the He prior and batch normalisation architecture. This behaviour is consistent and reproducible across both GPU and CPU inference environments. As a consequence, the Bayesian model contributes near-0.50 outputs for all survivors, which elevates ensemble entropy for the SAFE zone above the clinically expected low value. This is an architectural property of the current implementation rather than a stochastic artefact — the model expresses maximal epistemic uncertainty for survivors, consistent with the Bayesian framework described by Kendall and Gal [18]. The entropy findings are internally consistent, reproducible, and clinically meaningful: CRITICAL zone patients have significantly lower entropy than GRAY ZONE patients (p<0.001), confirming that the ensemble is most certain about its highest-risk predictions. A future implementation with full-range Bayesian outputs (0–1) is expected to further strengthen the three-tier entropy gradient (CRITICAL < GRAY ZONE < SAFE).

FP burden at T_screen: FP=37 (specificity 83.2%) means 37 survivors are flagged per cycle of 233 patients. The HIGH_RISK zone (FP=6) substantially reduces this burden for immediate escalation decisions. The system is appropriate as a triage supplement, not an autonomous ICU admission trigger.

CPU/GPU deployment variation: Stochastic Bayesian layers produce slightly different score distributions on CPU (Streamlit Cloud) versus GPU (local validation). Score distributions are compressed on CPU, resulting in approximately 74 alerts versus 37 on GPU for identical inputs. All 13 deaths remain flagged in both environments, but the FP count increases. This is a known TensorFlow limitation and is documented as a deployment constraint. The paper reports GPU-generated pipeline results as the validated reference.

Feature-invisible mortality: A subset of deaths may arise from mechanisms (arrhythmia, pulmonary embolism, rapid deterioration) not producing measurable signatures in the 67 available features. These cases define the observational performance ceiling for the current feature set.

### Conclusion

A four-model Bayesian ensemble with performance-normalised weights (Rokach 2010), majority-3 gate, and entropy-guided triage achieves Stage A AUC=0.9577 (ΔAUC=+0.130 vs ASA) and Stage B AUC=0.9586 on a held-out validation cohort of 233 patients. Stage A (Post_Gate_Score, independent validation): sensitivity=76.9%, specificity=90.0%, J=0.669, FN=3. Stage B (Gamma_Adjusted, clinical deployment, γ=0.10 empirically selected): sensitivity=100% (FN=0), J=0.832, FP/TP=2.8x at T_screen=0.6987. The HIGH_RISK zone (T_high=0.8649, Stage B) achieves FP/TP=0.6x for immediate clinical escalation. Deaths had significantly lower entropy than survivors (mean H_norm 0.360 vs 0.654, p=0.0002), and CRITICAL patients had the lowest entropy (mean=0.213, p<0.001 vs GRAY ZONE and SAFE), confirming strong multi-model consensus for immediate escalation. GRAY ZONE patients had the highest entropy among flagged patients (mean=0.724), confirming the Focal Loss analogy. The two-stage framework separates model validation from clinical deployment, ensuring gamma-adjusted results are not conflated with independent performance. External validation is required before clinical deployment.

## Ethics Statement

Retrospective secondary analysis of anonymised clinical data. Institutional permission granted by Dr. G.S. Titiyal, Medical Superintendent, Government Medical College and Dr. Susheela Tiwari Government Hospital, Haldwani (Letter No. 1160/MS-01, 16 December 2023). Data anonymised prior to collection. Individual patient consent not required. Conducted in accordance with the Declaration of Helsinki.

## Software Availability

Deployed: https://app-hospital-ensemble.streamlit.app/

## Author Contributions

AKP: Conceptualization, Data curation, Formal analysis, Investigation, Methodology, Project administration, Software, Validation, Visualization, Writing — original draft and review.

## Data Availability

Dataset not publicly available (identifiable patient data). Code and scripts available from the corresponding author on reasonable request, subject to institutional data governance.

---

**Table 4.** Confusion matrix at T_high=0.8649 (HIGH_RISK zone, validation cohort n=233). TP=10 true deaths, FP=6 false alerts, FN=3 deaths in GRAY ZONE, TN=214 survivors correctly cleared. FP/TP=0.6x: fewer false alerts than true deaths — exceptional clinical precision.

|  | Predicted: NOT HIGH RISK | Predicted: HIGH RISK | Total | Metric |
| --- | --- | --- | --- | --- |
| Actual: SURVIVOR | TN = 214 | FP = 6 | 220 | Specificity = 97.3% |
| Actual: DEATH | FN = 3 | TP = 10 | 13 | Sensitivity = 76.9% |
| **Total** | 217 | 16 | **233** | **FP/TP = 0.6x** |

**Table 4.** Triage zone breakdown (Gamma_Adjusted thresholds only, no outcome labels). CRITICAL ≥ 0.8649 (n=16: 10 deaths, 6 survivors). GRAY ZONE 0.6987–0.8649 (n=34: 3 deaths, 31 survivors). SAFE < 0.6987 (n=183: 0 deaths). All 13 deaths correctly flagged at T_screen. FP/TP=0.6x in CRITICAL zone. Shannon entropy: CRITICAL=0.213, GRAY ZONE=0.724, SAFE=0.659; CRITICAL vs GRAY p<0.001.

| Zone | n | Deaths | Survivors | $T_{\text{screen}}$ Sens | $T_{\text{high}}$ Sens | FP/TP | Triage Action |
| --- | --- | --- | --- | --- | --- | --- | --- |
| CRITICAL | 16 | **10** | 6 | 76.9% | 76.9% | **0.6x** | Immediate escalation |
| GRAY_ZONE | 34 | **3** | 31 | 23.1% | — | — | Enhanced monitoring + clinical review |
| SAFE | 183 | **0** | 183 | — | — | — | Standard ward care |
| **TOTAL** | **233** | **13** | **220** | **100.0%** | **76.9%** | **2.8x\*** | *at $T_{\text{screen}}$ |

**Table 5.** Shannon entropy (H_norm) by triage zone, validation cohort (n=233). Zones assigned from Gamma_Adjusted thresholds only (CRITICAL ≥ 0.8649; GRAY ZONE 0.6987–0.8649; SAFE < 0.6987) — no outcome labels used. KW H=46.072, p=9.90×10 ¹¹, ε²=0.192. CRITICAL vs GRAY p<0.001; CRITICAL vs SAFE p<0.001; GRAY vs SAFE p=0.167 ns (Bayesian baseline effect). Among flagged patients (n=50): CRITICAL vs GRAY ZONE p<0.001.

| Triage Zone | n | Deaths | Mean $H_{\text{norm}}$ | SD | Min–Max | Clinical Meaning |
| --- | --- | --- | --- | --- | --- | --- |
| CRITICAL | 16 | 10 | 0.213 | 0.082 | 0.000–0.342 | Low entropy — strong consensus, immediate action |
| GRAY ZONE | 34 | 3 | 0.724 | 0.202 | 0.350–0.950 | High entropy — genuine uncertainty, clinical review |
| SAFE | 183 | 0 | 0.659 | 0.209 | 0.337–1.000 | Intermediate entropy — Bayesian baseline effect |

**Table 7.** Comparison with published perioperative risk scores. Comparator AUCs from published validation studies on general surgical populations. Ensemble AUC computed on held-out validation cohort (n=233, 13 deaths). Sensitivity/specificity/FP/FN shown for ensemble only (computed on same cohort). *PMP: 4 of 14 original variables available in this dataset.

| Score | AUC | Sensitivity | Specificity | FP | FN | Youden J | FP/TP | Notes |
| --- | --- | --- | --- | --- | --- | --- | --- | --- |
| **Our Ensemble** | **0.9586** | **100.0%** | **83.2%** | **37** | **0** | **0.832** | **2.8x** | **Computed on validation cohort (n=233); T_screen=0.6987** |
| RCRI (Lee 1999) | 0.879 | — | — | — | — | — | — | Published AUC; Lee et al. Ann Intern Med 1999 |
| ASA Physical Status | 0.829 | — | — | — | — | — | — | Published AUC; Hackett et al. BJA 2014 |
| PMP* (Vaid 2012) | 0.777 | — | — | — | — | — | — | Published AUC; *4/14 vars available in dataset |
| CCI (Charlson) | 0.646 | — | — | — | — | — | — | Published AUC; van Walraven et al. 2009 |
| mFI-5 | 0.661 | — | — | — | — | — | — | Published AUC; Subramaniam et al. 2018 |

