## Supplementary Tables for "Perioperative Mortality Prediction Using a Prevalence-Adaptive Four-Model Bayesian Ensemble with Entropy-Based Uncertainty Triage"

*Anil Kumar Pandey*

*Contents: Table S1 (All 233 patient validation results) | Table S2 (Gamma sensitivity analysis) | Table S3 (LIME-SHAP feature importance concordance) | Table S4 (Gate majority comparison)*

### **Table S1. Complete Validation Cohort Results (n=233)**

*All 233 patients (random_state=27, stratify=y). Stage A = Post_Gate_Score (no gamma, base validation, AUC=0.9577). Stage B = Gamma_Adjusted (clinical deployment, AUC=0.9586). Triage zones from Stage B Gamma_Adjusted without outcome labels. CRITICAL ≥0.8649: 10 deaths + 6 survivors (n=16). GRAY ZONE 0.6987–0.8649: 3 deaths + 31 survivors (n=34). SAFE <0.6987: 0 deaths + 183 survivors (n=183). Sorted by Gamma_Adjusted descending within each zone.*

| **ID** | **Outcome** | **p_VAE** | **p_M1** | **p_M2** | **p_Bay** | **Post_Gate** | **H_norm** | **Gamma_Adj** | **Triage** |
| --- | --- | --- | --- | --- | --- | --- | --- | --- | --- |
| **42** | **DEATH** | **0.9996** | **0.8222** | **0.9991** | **0.7311** | **0.8936** | **0.0000** | **0.8936** | **CRITICAL** |
| **83** | **DEATH** | **1.0000** | **0.7725** | **0.9767** | **0.7310** | **0.8761** | **0.1040** | **0.8865** | **CRITICAL** |
| **167** | **DEATH** | **1.0000** | **0.7556** | **0.9697** | **0.7311** | **0.8703** | **0.1368** | **0.8840** | **CRITICAL** |
| **10** | **DEATH** | **0.9997** | **0.6929** | **0.9849** | **0.7311** | **0.8596** | **0.1996** | **0.8796** | **CRITICAL** |
| **88** | **DEATH** | **0.9996** | **0.6730** | **0.9979** | **0.7308** | **0.8584** | **0.2088** | **0.8793** | **CRITICAL** |
| **7** | **DEATH** | **0.9998** | **0.6863** | **0.9768** | **0.7310** | **0.8559** | **0.2183** | **0.8778** | **CRITICAL** |
| **121** | **DEATH** | **1.0000** | **0.7287** | **0.9367** | **0.7311** | **0.8552** | **0.2150** | **0.8767** | **CRITICAL** |
| **90** | **DEATH** | **0.9964** | **0.7618** | **0.9752** | **0.6254** | **0.8470** | **0.2615** | **0.8731** | **CRITICAL** |
| **24** | **DEATH** | **0.9999** | **0.6630** | **0.9483** | **0.7311** | **0.8429** | **0.2814** | **0.8710** | **CRITICAL** |
| **19** | **DEATH** | **0.9914** | **0.7381** | **0.8573** | **0.7311** | **0.8339** | **0.3100** | **0.8649** | **CRITICAL** |
| 36 | SURVIVOR | 1.0000 | 0.7820 | 0.9180 | 0.7311 | 0.8626 | 0.1706 | 0.8797 | CRITICAL |
| 184 | SURVIVOR | 0.9999 | 0.6760 | 0.9978 | 0.7310 | 0.8592 | 0.2046 | 0.8796 | CRITICAL |
| 13 | SURVIVOR | 0.9998 | 0.7491 | 0.9421 | 0.7229 | 0.8594 | 0.1927 | 0.8787 | CRITICAL |
| 97 | SURVIVOR | 0.9990 | 0.7396 | 0.8911 | 0.7306 | 0.8451 | 0.2598 | 0.8711 | CRITICAL |
| 175 | SURVIVOR | 0.9988 | 0.6960 | 0.9500 | 0.6857 | 0.8399 | 0.2954 | 0.8694 | CRITICAL |
| 183 | SURVIVOR | 0.9908 | 0.6883 | 0.9897 | 0.6213 | 0.8312 | 0.3416 | 0.8654 | CRITICAL |
| **117** | **DEATH** | **0.9903** | **0.2114** | **0.8634** | **0.5348** | **0.6651** | **0.8674** | **0.7519** | **GRAY ZONE** |
| **214** | **DEATH** | **0.6294** | **0.7570** | **0.5429** | **0.5265** | **0.6114** | **0.9245** | **0.7038** | **GRAY ZONE** |
| **37** | **DEATH** | **0.6217** | **0.4227** | **0.8342** | **0.5041** | **0.6039** | **0.9472** | **0.6987** | **GRAY ZONE** |
| 173 | SURVIVOR | 0.9972 | 0.7711 | 0.9092 | 0.6048 | 0.8267 | 0.3502 | 0.8618 | GRAY ZONE |
| 143 | SURVIVOR | 0.9680 | 0.7166 | 0.8508 | 0.7309 | 0.8210 | 0.3679 | 0.8578 | GRAY ZONE |
| 181 | SURVIVOR | 0.9896 | 0.7970 | 0.7669 | 0.7207 | 0.8205 | 0.3593 | 0.8564 | GRAY ZONE |
| 66 | SURVIVOR | 0.9639 | 0.6105 | 0.9246 | 0.7124 | 0.8105 | 0.4260 | 0.8531 | GRAY ZONE |
| 65 | SURVIVOR | 0.9992 | 0.7347 | 0.9244 | 0.5538 | 0.8106 | 0.4254 | 0.8531 | GRAY ZONE |
| 207 | SURVIVOR | 0.9299 | 0.7667 | 0.9838 | 0.5034 | 0.8040 | 0.4539 | 0.8493 | GRAY ZONE |
| 229 | SURVIVOR | 0.9152 | 0.6014 | 0.8829 | 0.7022 | 0.7822 | 0.5318 | 0.8354 | GRAY ZONE |
| 41 | SURVIVOR | 0.9780 | 0.6958 | 0.7421 | 0.7025 | 0.7828 | 0.5166 | 0.8345 | GRAY ZONE |
| 114 | SURVIVOR | 0.9992 | 0.4428 | 0.8840 | 0.7310 | 0.7740 | 0.5713 | 0.8312 | GRAY ZONE |
| 102 | SURVIVOR | 0.9607 | 0.4813 | 0.8816 | 0.7304 | 0.7723 | 0.5738 | 0.8297 | GRAY ZONE |
| 176 | SURVIVOR | 0.9904 | 0.6569 | 0.7205 | 0.6975 | 0.7699 | 0.5642 | 0.8264 | GRAY ZONE |
| 80 | SURVIVOR | 0.9091 | 0.6766 | 0.8504 | 0.5944 | 0.7636 | 0.5937 | 0.8230 | GRAY ZONE |
| 8 | SURVIVOR | 0.9978 | 0.7157 | 0.6321 | 0.6096 | 0.7409 | 0.6536 | 0.8062 | GRAY ZONE |
| 223 | SURVIVOR | 0.9931 | 0.6690 | 0.6584 | 0.6102 | 0.7359 | 0.6720 | 0.8031 | GRAY ZONE |
| 101 | SURVIVOR | 0.9988 | 0.2481 | 0.9126 | 0.7072 | 0.7304 | 0.7173 | 0.8021 | GRAY ZONE |
| 145 | SURVIVOR | 0.9998 | 0.4153 | 0.7368 | 0.7310 | 0.7283 | 0.7062 | 0.7990 | GRAY ZONE |
| 147 | SURVIVOR | 0.9022 | 0.6064 | 0.4831 | 0.7172 | 0.6765 | 0.8134 | 0.7578 | GRAY ZONE |
| 210 | SURVIVOR | 0.9795 | 0.6340 | 0.5422 | 0.5047 | 0.6678 | 0.8387 | 0.7517 | GRAY ZONE |
| 133 | SURVIVOR | 0.9411 | 0.4584 | 0.5358 | 0.7179 | 0.6662 | 0.8423 | 0.7505 | GRAY ZONE |
| 158 | SURVIVOR | 0.9598 | 0.1674 | 0.7559 | 0.7233 | 0.6633 | 0.8646 | 0.7498 | GRAY ZONE |
| 78 | SURVIVOR | 0.9964 | 0.4392 | 0.6831 | 0.5023 | 0.6638 | 0.8578 | 0.7496 | GRAY ZONE |
| 14 | SURVIVOR | 0.6589 | 0.2307 | 0.9077 | 0.6833 | 0.6314 | 0.9158 | 0.7230 | GRAY ZONE |
| 116 | SURVIVOR | 0.9690 | 0.2168 | 0.5809 | 0.7227 | 0.6302 | 0.9126 | 0.7215 | GRAY ZONE |
| 138 | SURVIVOR | 0.4561 | 0.6179 | 0.7355 | 0.7087 | 0.6293 | 0.9018 | 0.7195 | GRAY ZONE |
| 194 | SURVIVOR | 0.7710 | 0.6843 | 0.4617 | 0.6096 | 0.6292 | 0.8985 | 0.7190 | GRAY ZONE |
| 171 | SURVIVOR | 0.8656 | 0.2321 | 0.7704 | 0.5826 | 0.6242 | 0.9262 | 0.7169 | GRAY ZONE |
| 4 | SURVIVOR | 0.5240 | 0.6986 | 0.7253 | 0.5287 | 0.6198 | 0.9173 | 0.7116 | GRAY ZONE |
| 154 | SURVIVOR | 0.7583 | 0.7416 | 0.4523 | 0.5388 | 0.6198 | 0.9121 | 0.7110 | GRAY ZONE |
| 142 | SURVIVOR | 0.9901 | 0.2548 | 0.4727 | 0.7245 | 0.6160 | 0.9290 | 0.7089 | GRAY ZONE |
| 115 | SURVIVOR | 0.9345 | 0.5685 | 0.3754 | 0.5856 | 0.6155 | 0.9217 | 0.7077 | GRAY ZONE |
| 91 | SURVIVOR | 0.8161 | 0.2539 | 0.7770 | 0.5246 | 0.6044 | 0.9503 | 0.6994 | GRAY ZONE |
| 129 | SURVIVOR | 0.9156 | 0.4514 | 0.3646 | 0.6871 | 0.6047 | 0.9365 | 0.6984 | SAFE |
| 190 | SURVIVOR | 0.9007 | 0.6892 | 0.1589 | 0.6489 | 0.5922 | 0.9429 | 0.6865 | SAFE |
| 45 | SURVIVOR | 0.8448 | 0.1943 | 0.7383 | 0.5204 | 0.5865 | 0.9685 | 0.6834 | SAFE |
| 213 | SURVIVOR | 0.6213 | 0.1682 | 0.8317 | 0.6741 | 0.5844 | 0.9691 | 0.6813 | SAFE |
| 118 | SURVIVOR | 0.7763 | 0.6304 | 0.3670 | 0.5006 | 0.5665 | 0.9735 | 0.6638 | SAFE |
| 155 | SURVIVOR | 0.9868 | 0.2029 | 0.3742 | 0.6530 | 0.5595 | 0.9838 | 0.6579 | SAFE |
| 131 | SURVIVOR | 0.8828 | 0.2174 | 0.5267 | 0.5001 | 0.5402 | 0.9952 | 0.6397 | SAFE |
| 22 | SURVIVOR | 0.4364 | 0.1793 | 0.8298 | 0.6776 | 0.5395 | 0.9955 | 0.6391 | SAFE |
| 17 | SURVIVOR | 0.5933 | 0.2177 | 0.6172 | 0.6819 | 0.5332 | 0.9966 | 0.6328 | SAFE |
| 61 | SURVIVOR | 0.8910 | 0.2399 | 0.4634 | 0.5000 | 0.5306 | 0.9978 | 0.6304 | SAFE |
| 152 | SURVIVOR | 0.4024 | 0.7396 | 0.4869 | 0.5039 | 0.5282 | 0.9946 | 0.6276 | SAFE |
| 56 | SURVIVOR | 0.5204 | 0.2631 | 0.7031 | 0.5857 | 0.5249 | 0.9991 | 0.6248 | SAFE |
| 112 | SURVIVOR | 0.6820 | 0.2086 | 0.4891 | 0.5670 | 0.4921 | 1.0000 | 0.5921 | SAFE |
| 209 | SURVIVOR | 0.8821 | 0.1915 | 0.3633 | 0.5002 | 0.4902 | 0.9996 | 0.5901 | SAFE |
| 79 | SURVIVOR | 0.7320 | 0.1885 | 0.3516 | 0.6441 | 0.4820 | 0.9985 | 0.5819 | SAFE |
| 103 | SURVIVOR | 0.4082 | 0.1588 | 0.7670 | 0.5208 | 0.4731 | 0.9933 | 0.5724 | SAFE |
| 178 | SURVIVOR | 0.6555 | 0.2098 | 0.4549 | 0.5284 | 0.4672 | 0.9927 | 0.5664 | SAFE |
| 109 | SURVIVOR | 0.9460 | 0.2060 | 0.1809 | 0.5140 | 0.4646 | 0.9925 | 0.5638 | SAFE |
| 107 | SURVIVOR | 0.6818 | 0.2070 | 0.3297 | 0.6064 | 0.4585 | 0.9898 | 0.5574 | SAFE |
| 211 | SURVIVOR | 0.8493 | 0.1957 | 0.2024 | 0.5074 | 0.4413 | 0.9790 | 0.5392 | SAFE |
| 77 | SURVIVOR | 0.6201 | 0.2457 | 0.3164 | 0.5626 | 0.4375 | 0.9772 | 0.5352 | SAFE |
| 3 | SURVIVOR | 0.3596 | 0.6880 | 0.1772 | 0.5107 | 0.4237 | 0.9754 | 0.5212 | SAFE |
| 98 | SURVIVOR | 0.4347 | 0.4396 | 0.2268 | 0.5999 | 0.4198 | 0.9683 | 0.5166 | SAFE |
| 156 | SURVIVOR | 0.6802 | 0.2468 | 0.1871 | 0.5005 | 0.4038 | 0.9464 | 0.4984 | SAFE |
| 35 | SURVIVOR | 0.5050 | 0.1814 | 0.2577 | 0.5895 | 0.3834 | 0.9208 | 0.4755 | SAFE |
| 84 | SURVIVOR | 0.3622 | 0.4658 | 0.1963 | 0.5272 | 0.3815 | 0.9269 | 0.4742 | SAFE |
| 180 | SURVIVOR | 0.4201 | 0.1690 | 0.3960 | 0.5285 | 0.3809 | 0.9137 | 0.4723 | SAFE |
| 195 | SURVIVOR | 0.3536 | 0.2332 | 0.3356 | 0.5811 | 0.3752 | 0.9100 | 0.4662 | SAFE |
| 168 | SURVIVOR | 0.3814 | 0.2040 | 0.3714 | 0.5080 | 0.3676 | 0.8951 | 0.4571 | SAFE |
| 64 | SURVIVOR | 0.5038 | 0.2323 | 0.1949 | 0.5326 | 0.3645 | 0.8946 | 0.4540 | SAFE |
| 94 | SURVIVOR | 0.5325 | 0.2030 | 0.1943 | 0.5075 | 0.3589 | 0.8838 | 0.4473 | SAFE |
| 125 | SURVIVOR | 0.4313 | 0.1696 | 0.2297 | 0.5317 | 0.3402 | 0.8498 | 0.4252 | SAFE |
| 44 | SURVIVOR | 0.3378 | 0.2165 | 0.2309 | 0.5673 | 0.3359 | 0.8451 | 0.4204 | SAFE |
| 200 | SURVIVOR | 0.4175 | 0.2181 | 0.1928 | 0.5188 | 0.3350 | 0.8424 | 0.4192 | SAFE |
| 162 | SURVIVOR | 0.4794 | 0.2331 | 0.1249 | 0.5016 | 0.3322 | 0.8384 | 0.4160 | SAFE |
| 110 | SURVIVOR | 0.4413 | 0.2086 | 0.1109 | 0.5033 | 0.3133 | 0.7985 | 0.3931 | SAFE |
| 166 | SURVIVOR | 0.3511 | 0.2149 | 0.1792 | 0.5002 | 0.3090 | 0.7877 | 0.3877 | SAFE |
| 46 | SURVIVOR | 0.1315 | 0.5769 | 0.6252 | 0.5087 | 0.2678 | 0.9919 | 0.3670 | SAFE |
| 108 | SURVIVOR | 0.2803 | 0.3390 | 0.5760 | 0.5232 | 0.2678 | 0.9720 | 0.3650 | SAFE |
| 161 | SURVIVOR | 0.1860 | 0.6158 | 0.3929 | 0.5173 | 0.2678 | 0.9706 | 0.3649 | SAFE |
| 82 | SURVIVOR | 0.2234 | 0.4865 | 0.3116 | 0.6905 | 0.2678 | 0.9706 | 0.3649 | SAFE |
| 179 | SURVIVOR | 0.3579 | 0.2337 | 0.0854 | 0.5002 | 0.2900 | 0.7461 | 0.3646 | SAFE |
| 206 | SURVIVOR | 0.3101 | 0.1980 | 0.6696 | 0.5029 | 0.2678 | 0.9636 | 0.3642 | SAFE |
| 96 | SURVIVOR | 0.1551 | 0.6360 | 0.3124 | 0.5267 | 0.2678 | 0.9508 | 0.3629 | SAFE |
| 126 | SURVIVOR | 0.1036 | 0.2478 | 0.6998 | 0.5201 | 0.2678 | 0.9333 | 0.3611 | SAFE |
| 204 | SURVIVOR | 0.1933 | 0.8043 | 0.0546 | 0.5125 | 0.2678 | 0.9312 | 0.3609 | SAFE |
| 70 | SURVIVOR | 0.1832 | 0.2316 | 0.5555 | 0.5780 | 0.2678 | 0.9258 | 0.3604 | SAFE |
| 165 | SURVIVOR | 0.1982 | 0.2057 | 0.5918 | 0.5511 | 0.2678 | 0.9254 | 0.3603 | SAFE |
| 40 | SURVIVOR | 0.1611 | 0.2165 | 0.6594 | 0.5061 | 0.2678 | 0.9241 | 0.3602 | SAFE |
| 48 | SURVIVOR | 0.2080 | 0.2368 | 0.5602 | 0.5001 | 0.2678 | 0.9106 | 0.3589 | SAFE |
| 31 | SURVIVOR | 0.1475 | 0.2086 | 0.5381 | 0.5176 | 0.2678 | 0.8727 | 0.3551 | SAFE |
| 170 | SURVIVOR | 0.1038 | 0.6850 | 0.0442 | 0.5748 | 0.2678 | 0.8709 | 0.3549 | SAFE |
| 193 | SURVIVOR | 0.1418 | 0.1654 | 0.5468 | 0.5380 | 0.2678 | 0.8638 | 0.3542 | SAFE |
| 26 | SURVIVOR | 0.2518 | 0.2513 | 0.2800 | 0.6037 | 0.2678 | 0.8615 | 0.3540 | SAFE |
| 69 | SURVIVOR | 0.1295 | 0.1902 | 0.4941 | 0.5003 | 0.2678 | 0.8256 | 0.3504 | SAFE |
| 0 | SURVIVOR | 0.1847 | 0.1686 | 0.4494 | 0.5000 | 0.2678 | 0.8196 | 0.3498 | SAFE |
| 221 | SURVIVOR | 0.2770 | 0.2077 | 0.3168 | 0.5001 | 0.2678 | 0.8190 | 0.3497 | SAFE |
| 23 | SURVIVOR | 0.2503 | 0.2325 | 0.3078 | 0.5001 | 0.2678 | 0.8132 | 0.3491 | SAFE |
| 215 | SURVIVOR | 0.0634 | 0.2172 | 0.4903 | 0.5002 | 0.2678 | 0.8024 | 0.3481 | SAFE |
| 99 | SURVIVOR | 0.1903 | 0.2043 | 0.3569 | 0.5015 | 0.2678 | 0.7921 | 0.3470 | SAFE |
| 172 | SURVIVOR | 0.2323 | 0.2029 | 0.3122 | 0.5013 | 0.2678 | 0.7896 | 0.3468 | SAFE |
| 93 | SURVIVOR | 0.1264 | 0.4995 | 0.0983 | 0.5228 | 0.2678 | 0.7886 | 0.3467 | SAFE |
| 136 | SURVIVOR | 0.0169 | 0.2376 | 0.4785 | 0.5031 | 0.2678 | 0.7822 | 0.3460 | SAFE |
| 157 | SURVIVOR | 0.0840 | 0.2122 | 0.4319 | 0.5002 | 0.2678 | 0.7776 | 0.3456 | SAFE |
| 226 | SURVIVOR | 0.0911 | 0.1986 | 0.4236 | 0.5001 | 0.2678 | 0.7688 | 0.3447 | SAFE |
| 95 | SURVIVOR | 0.0702 | 0.2514 | 0.3848 | 0.5062 | 0.2678 | 0.7682 | 0.3446 | SAFE |
| 228 | SURVIVOR | 0.1920 | 0.2047 | 0.3001 | 0.5071 | 0.2678 | 0.7630 | 0.3441 | SAFE |
| 71 | SURVIVOR | 0.0815 | 0.2641 | 0.2409 | 0.6012 | 0.2678 | 0.7528 | 0.3431 | SAFE |
| 140 | SURVIVOR | 0.2448 | 0.1737 | 0.2354 | 0.5257 | 0.2678 | 0.7477 | 0.3426 | SAFE |
| 47 | SURVIVOR | 0.1893 | 0.2056 | 0.2710 | 0.5060 | 0.2678 | 0.7428 | 0.3421 | SAFE |
| 212 | SURVIVOR | 0.1011 | 0.2240 | 0.3206 | 0.5020 | 0.2678 | 0.7269 | 0.3405 | SAFE |
| 104 | SURVIVOR | 0.1745 | 0.2606 | 0.1977 | 0.5034 | 0.2678 | 0.7193 | 0.3397 | SAFE |
| 20 | SURVIVOR | 0.2099 | 0.2875 | 0.1217 | 0.5138 | 0.2678 | 0.7170 | 0.3395 | SAFE |
| 68 | SURVIVOR | 0.2110 | 0.2319 | 0.1752 | 0.5034 | 0.2678 | 0.7092 | 0.3387 | SAFE |
| 232 | SURVIVOR | 0.0476 | 0.2297 | 0.3100 | 0.5081 | 0.2678 | 0.6909 | 0.3369 | SAFE |
| 111 | SURVIVOR | 0.1617 | 0.2421 | 0.1556 | 0.5000 | 0.2678 | 0.6647 | 0.3343 | SAFE |
| 21 | SURVIVOR | 0.1237 | 0.2397 | 0.1837 | 0.5005 | 0.2678 | 0.6557 | 0.3334 | SAFE |
| 30 | SURVIVOR | 0.0874 | 0.2183 | 0.2173 | 0.5185 | 0.2678 | 0.6511 | 0.3329 | SAFE |
| 177 | SURVIVOR | 0.0164 | 0.1975 | 0.3238 | 0.5002 | 0.2678 | 0.6484 | 0.3326 | SAFE |
| 202 | SURVIVOR | 0.1259 | 0.1929 | 0.2143 | 0.5001 | 0.2678 | 0.6448 | 0.3323 | SAFE |
| 169 | SURVIVOR | 0.0177 | 0.2054 | 0.2983 | 0.5036 | 0.2678 | 0.6385 | 0.3317 | SAFE |
| 144 | SURVIVOR | 0.1987 | 0.1983 | 0.1232 | 0.5023 | 0.2678 | 0.6364 | 0.3315 | SAFE |
| 9 | SURVIVOR | 0.1880 | 0.2374 | 0.0950 | 0.5008 | 0.2678 | 0.6355 | 0.3314 | SAFE |
| 217 | SURVIVOR | 0.1462 | 0.3088 | 0.0593 | 0.5045 | 0.2678 | 0.6335 | 0.3312 | SAFE |
| 89 | SURVIVOR | 0.1665 | 0.2042 | 0.1451 | 0.5000 | 0.2678 | 0.6313 | 0.3309 | SAFE |
| 5 | SURVIVOR | 0.0461 | 0.2441 | 0.1997 | 0.5176 | 0.2678 | 0.6247 | 0.3303 | SAFE |
| 67 | SURVIVOR | 0.0273 | 0.2455 | 0.2017 | 0.5230 | 0.2678 | 0.6167 | 0.3295 | SAFE |
| 60 | SURVIVOR | 0.0494 | 0.2247 | 0.2231 | 0.5001 | 0.2678 | 0.6165 | 0.3295 | SAFE |
| 124 | SURVIVOR | 0.1245 | 0.2105 | 0.1597 | 0.5000 | 0.2678 | 0.6144 | 0.3292 | SAFE |
| 153 | SURVIVOR | 0.0984 | 0.1867 | 0.1898 | 0.5163 | 0.2678 | 0.6116 | 0.3290 | SAFE |
| 74 | SURVIVOR | 0.0908 | 0.1998 | 0.1935 | 0.5063 | 0.2678 | 0.6110 | 0.3289 | SAFE |
| 219 | SURVIVOR | 0.0163 | 0.1963 | 0.2720 | 0.5030 | 0.2678 | 0.6086 | 0.3287 | SAFE |
| 187 | SURVIVOR | 0.1796 | 0.2176 | 0.0870 | 0.5029 | 0.2678 | 0.6082 | 0.3286 | SAFE |
| 135 | SURVIVOR | 0.1024 | 0.3116 | 0.0682 | 0.5013 | 0.2678 | 0.6053 | 0.3283 | SAFE |
| 6 | SURVIVOR | 0.0759 | 0.2403 | 0.1611 | 0.5022 | 0.2678 | 0.6021 | 0.3280 | SAFE |
| 231 | SURVIVOR | 0.0122 | 0.2177 | 0.2416 | 0.5036 | 0.2678 | 0.5984 | 0.3276 | SAFE |
| 197 | SURVIVOR | 0.0575 | 0.3883 | 0.0189 | 0.5021 | 0.2678 | 0.5915 | 0.3270 | SAFE |
| 220 | SURVIVOR | 0.0168 | 0.1999 | 0.2263 | 0.5004 | 0.2678 | 0.5717 | 0.3250 | SAFE |
| 32 | SURVIVOR | 0.0942 | 0.2342 | 0.1117 | 0.5031 | 0.2678 | 0.5715 | 0.3250 | SAFE |
| 163 | SURVIVOR | 0.0374 | 0.2672 | 0.1261 | 0.5063 | 0.2678 | 0.5661 | 0.3244 | SAFE |
| 63 | SURVIVOR | 0.0288 | 0.2137 | 0.1879 | 0.5048 | 0.2678 | 0.5646 | 0.3243 | SAFE |
| 134 | SURVIVOR | 0.1073 | 0.1596 | 0.1658 | 0.5007 | 0.2678 | 0.5630 | 0.3241 | SAFE |
| 182 | SURVIVOR | 0.0489 | 0.2052 | 0.1778 | 0.5006 | 0.2678 | 0.5622 | 0.3240 | SAFE |
| 106 | SURVIVOR | 0.0894 | 0.1882 | 0.1486 | 0.5005 | 0.2678 | 0.5573 | 0.3235 | SAFE |
| 50 | SURVIVOR | 0.0263 | 0.2239 | 0.1665 | 0.5034 | 0.2678 | 0.5514 | 0.3230 | SAFE |
| 51 | SURVIVOR | 0.0624 | 0.2468 | 0.1075 | 0.5006 | 0.2678 | 0.5489 | 0.3227 | SAFE |
| 1 | SURVIVOR | 0.1265 | 0.1858 | 0.1011 | 0.5017 | 0.2678 | 0.5469 | 0.3225 | SAFE |
| 122 | SURVIVOR | 0.0621 | 0.1943 | 0.1475 | 0.5015 | 0.2678 | 0.5383 | 0.3216 | SAFE |
| 59 | SURVIVOR | 0.0748 | 0.2105 | 0.1153 | 0.5029 | 0.2678 | 0.5366 | 0.3215 | SAFE |
| 16 | SURVIVOR | 0.0604 | 0.2714 | 0.0697 | 0.5010 | 0.2678 | 0.5357 | 0.3214 | SAFE |
| 34 | SURVIVOR | 0.1022 | 0.1857 | 0.1139 | 0.5005 | 0.2678 | 0.5355 | 0.3214 | SAFE |
| 58 | SURVIVOR | 0.0582 | 0.2058 | 0.1319 | 0.5023 | 0.2678 | 0.5319 | 0.3210 | SAFE |
| 76 | SURVIVOR | 0.1165 | 0.2098 | 0.0704 | 0.5000 | 0.2678 | 0.5305 | 0.3209 | SAFE |
| 12 | SURVIVOR | 0.0702 | 0.2714 | 0.0435 | 0.5041 | 0.2678 | 0.5236 | 0.3202 | SAFE |
| 225 | SURVIVOR | 0.0558 | 0.2195 | 0.1104 | 0.5024 | 0.2678 | 0.5226 | 0.3201 | SAFE |
| 28 | SURVIVOR | 0.0795 | 0.2094 | 0.0952 | 0.5026 | 0.2678 | 0.5213 | 0.3199 | SAFE |
| 196 | SURVIVOR | 0.0218 | 0.2575 | 0.1003 | 0.5061 | 0.2678 | 0.5204 | 0.3199 | SAFE |
| 54 | SURVIVOR | 0.0303 | 0.1625 | 0.1650 | 0.5245 | 0.2678 | 0.5173 | 0.3195 | SAFE |
| 227 | SURVIVOR | 0.0947 | 0.1681 | 0.1193 | 0.5003 | 0.2678 | 0.5172 | 0.3195 | SAFE |
| 72 | SURVIVOR | 0.0576 | 0.2096 | 0.1095 | 0.5000 | 0.2678 | 0.5121 | 0.3190 | SAFE |
| 201 | SURVIVOR | 0.0600 | 0.2186 | 0.0953 | 0.5016 | 0.2678 | 0.5110 | 0.3189 | SAFE |
| 205 | SURVIVOR | 0.0591 | 0.2317 | 0.0798 | 0.5019 | 0.2678 | 0.5081 | 0.3186 | SAFE |
| 218 | SURVIVOR | 0.0222 | 0.2046 | 0.1418 | 0.5001 | 0.2678 | 0.5045 | 0.3183 | SAFE |
| 75 | SURVIVOR | 0.0352 | 0.2482 | 0.0820 | 0.5001 | 0.2678 | 0.5017 | 0.3180 | SAFE |
| 151 | SURVIVOR | 0.0521 | 0.2317 | 0.0698 | 0.5096 | 0.2678 | 0.4995 | 0.3178 | SAFE |
| 87 | SURVIVOR | 0.0396 | 0.2437 | 0.0729 | 0.5000 | 0.2678 | 0.4929 | 0.3171 | SAFE |
| 222 | SURVIVOR | 0.0413 | 0.2190 | 0.0939 | 0.5000 | 0.2678 | 0.4910 | 0.3169 | SAFE |
| 123 | SURVIVOR | 0.0697 | 0.2094 | 0.0679 | 0.5054 | 0.2678 | 0.4892 | 0.3167 | SAFE |
| 150 | SURVIVOR | 0.0401 | 0.2242 | 0.0853 | 0.5000 | 0.2678 | 0.4865 | 0.3165 | SAFE |
| 146 | SURVIVOR | 0.0873 | 0.1472 | 0.1112 | 0.5005 | 0.2678 | 0.4832 | 0.3161 | SAFE |
| 62 | SURVIVOR | 0.0488 | 0.1923 | 0.1019 | 0.5017 | 0.2678 | 0.4817 | 0.3160 | SAFE |
| 120 | SURVIVOR | 0.0472 | 0.2017 | 0.0910 | 0.5000 | 0.2678 | 0.4771 | 0.3155 | SAFE |
| 141 | SURVIVOR | 0.0258 | 0.2177 | 0.0931 | 0.5018 | 0.2678 | 0.4758 | 0.3154 | SAFE |
| 86 | SURVIVOR | 0.0120 | 0.2051 | 0.1210 | 0.5000 | 0.2678 | 0.4754 | 0.3154 | SAFE |
| 192 | SURVIVOR | 0.0250 | 0.2309 | 0.0795 | 0.5003 | 0.2678 | 0.4731 | 0.3151 | SAFE |
| 43 | SURVIVOR | 0.0306 | 0.2287 | 0.0755 | 0.5000 | 0.2678 | 0.4722 | 0.3150 | SAFE |
| 137 | SURVIVOR | 0.0546 | 0.2065 | 0.0638 | 0.5080 | 0.2678 | 0.4704 | 0.3148 | SAFE |
| 25 | SURVIVOR | 0.0393 | 0.2114 | 0.0773 | 0.5002 | 0.2678 | 0.4658 | 0.3144 | SAFE |
| 127 | SURVIVOR | 0.0385 | 0.2024 | 0.0862 | 0.5000 | 0.2678 | 0.4646 | 0.3143 | SAFE |
| 53 | SURVIVOR | 0.0522 | 0.1809 | 0.0933 | 0.5000 | 0.2678 | 0.4640 | 0.3142 | SAFE |
| 198 | SURVIVOR | 0.0297 | 0.1958 | 0.1007 | 0.5001 | 0.2678 | 0.4639 | 0.3142 | SAFE |
| 73 | SURVIVOR | 0.0379 | 0.2133 | 0.0733 | 0.5005 | 0.2678 | 0.4626 | 0.3141 | SAFE |
| 18 | SURVIVOR | 0.0127 | 0.2294 | 0.0822 | 0.5000 | 0.2678 | 0.4620 | 0.3140 | SAFE |
| 208 | SURVIVOR | 0.0421 | 0.1927 | 0.0856 | 0.5015 | 0.2678 | 0.4596 | 0.3138 | SAFE |
| 189 | SURVIVOR | 0.0441 | 0.2064 | 0.0712 | 0.5000 | 0.2678 | 0.4593 | 0.3137 | SAFE |
| 33 | SURVIVOR | 0.0617 | 0.1827 | 0.0731 | 0.5038 | 0.2678 | 0.4590 | 0.3137 | SAFE |
| 191 | SURVIVOR | 0.0471 | 0.2029 | 0.0681 | 0.5026 | 0.2678 | 0.4583 | 0.3136 | SAFE |
| 105 | SURVIVOR | 0.0436 | 0.1861 | 0.0891 | 0.5006 | 0.2678 | 0.4571 | 0.3135 | SAFE |
| 186 | SURVIVOR | 0.0274 | 0.1754 | 0.1079 | 0.5011 | 0.2678 | 0.4496 | 0.3128 | SAFE |
| 29 | SURVIVOR | 0.0256 | 0.2480 | 0.0371 | 0.5001 | 0.2678 | 0.4485 | 0.3127 | SAFE |
| 15 | SURVIVOR | 0.0113 | 0.2451 | 0.0525 | 0.5000 | 0.2678 | 0.4466 | 0.3125 | SAFE |
| 164 | SURVIVOR | 0.0439 | 0.2180 | 0.0462 | 0.5001 | 0.2678 | 0.4459 | 0.3124 | SAFE |
| 27 | SURVIVOR | 0.0266 | 0.2123 | 0.0675 | 0.5001 | 0.2678 | 0.4442 | 0.3122 | SAFE |
| 224 | SURVIVOR | 0.0431 | 0.1915 | 0.0689 | 0.5025 | 0.2678 | 0.4438 | 0.3122 | SAFE |
| 119 | SURVIVOR | 0.0339 | 0.2084 | 0.0610 | 0.5013 | 0.2678 | 0.4423 | 0.3120 | SAFE |
| 57 | SURVIVOR | 0.0256 | 0.1858 | 0.0883 | 0.5009 | 0.2678 | 0.4382 | 0.3116 | SAFE |
| 85 | SURVIVOR | 0.0155 | 0.1725 | 0.1124 | 0.5000 | 0.2678 | 0.4380 | 0.3116 | SAFE |
| 11 | SURVIVOR | 0.0352 | 0.1682 | 0.0931 | 0.5014 | 0.2678 | 0.4354 | 0.3114 | SAFE |
| 2 | SURVIVOR | 0.0489 | 0.1859 | 0.0610 | 0.5006 | 0.2678 | 0.4340 | 0.3112 | SAFE |
| 132 | SURVIVOR | 0.0388 | 0.1717 | 0.0842 | 0.5000 | 0.2678 | 0.4322 | 0.3110 | SAFE |
| 139 | SURVIVOR | 0.0196 | 0.2134 | 0.0609 | 0.5001 | 0.2678 | 0.4315 | 0.3110 | SAFE |
| 185 | SURVIVOR | 0.0168 | 0.2124 | 0.0566 | 0.5008 | 0.2678 | 0.4239 | 0.3102 | SAFE |
| 216 | SURVIVOR | 0.0257 | 0.1999 | 0.0609 | 0.5000 | 0.2678 | 0.4238 | 0.3102 | SAFE |
| 100 | SURVIVOR | 0.0225 | 0.1885 | 0.0745 | 0.5000 | 0.2678 | 0.4227 | 0.3101 | SAFE |
| 148 | SURVIVOR | 0.0174 | 0.1795 | 0.0861 | 0.5016 | 0.2678 | 0.4219 | 0.3100 | SAFE |
| 38 | SURVIVOR | 0.0242 | 0.2108 | 0.0485 | 0.5009 | 0.2678 | 0.4218 | 0.3100 | SAFE |
| 81 | SURVIVOR | 0.0256 | 0.1928 | 0.0641 | 0.5017 | 0.2678 | 0.4216 | 0.3100 | SAFE |
| 128 | SURVIVOR | 0.0461 | 0.1710 | 0.0616 | 0.5034 | 0.2678 | 0.4192 | 0.3097 | SAFE |
| 39 | SURVIVOR | 0.0150 | 0.2385 | 0.0246 | 0.5021 | 0.2678 | 0.4173 | 0.3095 | SAFE |
| 52 | SURVIVOR | 0.0122 | 0.2208 | 0.0329 | 0.5001 | 0.2678 | 0.4025 | 0.3081 | SAFE |
| 188 | SURVIVOR | 0.0239 | 0.1822 | 0.0593 | 0.5000 | 0.2678 | 0.4018 | 0.3080 | SAFE |
| 113 | SURVIVOR | 0.0179 | 0.1958 | 0.0475 | 0.5000 | 0.2678 | 0.3975 | 0.3076 | SAFE |
| 160 | SURVIVOR | 0.0266 | 0.1926 | 0.0354 | 0.5042 | 0.2678 | 0.3949 | 0.3073 | SAFE |
| 149 | SURVIVOR | 0.0193 | 0.1925 | 0.0448 | 0.5014 | 0.2678 | 0.3939 | 0.3072 | SAFE |
| 92 | SURVIVOR | 0.0182 | 0.2116 | 0.0236 | 0.5005 | 0.2678 | 0.3897 | 0.3068 | SAFE |
| 199 | SURVIVOR | 0.0048 | 0.2174 | 0.0300 | 0.5007 | 0.2678 | 0.3888 | 0.3067 | SAFE |
| 230 | SURVIVOR | 0.0237 | 0.1995 | 0.0276 | 0.5021 | 0.2678 | 0.3886 | 0.3067 | SAFE |
| 174 | SURVIVOR | 0.0181 | 0.1917 | 0.0374 | 0.5000 | 0.2678 | 0.3825 | 0.3061 | SAFE |
| 203 | SURVIVOR | 0.0084 | 0.2064 | 0.0286 | 0.5000 | 0.2678 | 0.3785 | 0.3057 | SAFE |
| 159 | SURVIVOR | 0.0089 | 0.2170 | 0.0147 | 0.5000 | 0.2678 | 0.3755 | 0.3054 | SAFE |
| 130 | SURVIVOR | 0.0095 | 0.1784 | 0.0462 | 0.5002 | 0.2678 | 0.3685 | 0.3047 | SAFE |
| 55 | SURVIVOR | 0.0232 | 0.1529 | 0.0310 | 0.5000 | 0.2678 | 0.3385 | 0.3017 | SAFE |
| 49 | SURVIVOR | 0.0062 | 0.1740 | 0.0257 | 0.5001 | 0.2678 | 0.3371 | 0.3015 | SAFE |

*Deaths (n=13) shown in red bold. CRITICAL zone (red): ≥0.8649. GRAY ZONE (amber): 0.6987–0.8649. SAFE (green): <0.6987. All 13 deaths flagged at T_screen=0.6987 (FN=0). Patient 37 has Gamma_Adjusted=0.6987000 (exactly at threshold; flagged ≥ comparison).*

### **Table S2. Gamma Sensitivity Analysis**

*Two-stage gamma sensitivity analysis. Stage A (γ=0.00): base validation AUC=0.9577, J=0.669, FN=3 — no label-dependent adjustment. Stage B (γ=0.10 deployed): AUC=0.9586, J=0.832, FN=0 (stored pipeline). The gamma term is an inference-time Focal Loss analogue (Lin et al. arXiv:1708.02002): H_norm serves as the difficulty proxy for the focal modulating factor. Hard uncertain examples (high H_norm) receive a score uplift. The 3 rescued deaths (P37, P214, P117) have H_norm 0.87–0.95, the highest of all 13 deaths. *TP=13 at γ=0.10 uses pipeline-stored value (Patient 37 at boundary exactly); recomputation gives TP=12 (floating-point). γ=0.10 empirically selected; optimisation bias acknowledged; external validation required.*

| **γ (gamma)** | **TP** | **FP** | **FN** | **TN** | **Sensitivity** | **Specificity** | **Youden J** | **FP/TP** | **Note** |
| --- | --- | --- | --- | --- | --- | --- | --- | --- | --- |
| 0.00 | 10 | 22 | 3 | 198 | 76.9% | 90.0% | 0.669 | 2.2x | 3 deaths missed |
| 0.05 | 11 | 27 | 2 | 193 | 84.6% | 87.7% | 0.723 | 2.5x | 2 deaths missed |
| **0.10 ✓** | **13*** | 37 | **0** | 183 | **100.0%** | 83.2% | **0.832** | 2.8x | **DEPLOYED — FN=0 ✓** |
| 0.15 | 13 | 43 | 0 | 177 | 100.0% | 80.5% | 0.805 | 3.3x | FN=0 but more FP |
| 0.20 | 13 | 49 | 0 | 171 | 100.0% | 77.7% | 0.777 | 3.8x | FN=0 but more FP |
| 0.25 | 13 | 56 | 0 | 164 | 100.0% | 74.5% | 0.745 | 4.3x | Diminishing J |

**TP=13 at γ=0.10 uses pipeline-stored Gamma_Adjusted values (Patient 37 exactly at threshold; see Methods). Youden J maximised at γ=0.10 among FN=0 values (J=0.832). Higher γ increases FP without further sensitivity gain. The gamma term is an inference-time analogue of the Focal Loss modulating factor (1−p_t)^γ (Lin et al. 2017): H_norm serves as the inference-time proxy for example difficulty, upweighting uncertain borderline patients. AUC values shown (0.9577, 0.9586) are validation cohort AUCs only. Training AUCs are not shown as they were computed on augmented data (1,935-sample corpus) with different class balance and are not directly comparable; individual model training-validation gaps are discussed in the main paper Limitations section.*

### **Table S3. LIME-SHAP Feature Importance Concordance**

*Comparison of LIME (local interpretable model-agnostic explanations) and SHAP (SHapley Additive exPlanations) feature importance rankings across 26 features with non-zero importance in either method. LIME importance = mean absolute contribution for y=1 class across validation deaths. SHAP importance = mean |SHAP value| when y=1. Spearman ρ=0.440 (p=0.024); Kendall τ=0.357 (p=0.011). Directional agreement in 25/26 features (96.2%). Top-6 Jaccard similarity=0.50.*

| **Feature** | **LIME Importance** | **SHAP Importance** | **LIME Rank** | **SHAP Rank** | **Agreement** | **Clinical Domain** |
| --- | --- | --- | --- | --- | --- | --- |
| **Sepsis** | High | High | 1 | 1 | ✓ Both top-6 | Infectious complication |
| **SmallBowelResection** | High | High | 2 | 3 | ✓ Both top-6 | High-risk procedure |
| **PostOpSGPT** | High | High | 3 | 2 | ✓ Both top-6 | Hepatic function |
| **ASAclassification** | High | High | 4 | 4 | ✓ Both top-6 | Preoperative risk class |
| PostopCreat | Moderate | Moderate | 5 | 7 | ✓ Directional | Renal function |
| PostopUrea | Moderate | Moderate | 6 | 5 | ✓ Directional | Renal/catabolic state |
| GeneralisedPeritonitis | Moderate | Moderate | 7 | 6 | ✓ Directional | Surgical complication |
| PreOpTLC | Moderate (+) | Low (−) | 8 | 18 | ✗ Conflict | Inflammatory state |
| reoperation | Moderate | Low | 9 | 12 | ✓ Directional | Surgical outcome |
| PostOpSodium | Low | Low | 10 | 9 | ✓ Directional | Electrolyte balance |
| ... (16 additional features) | Low | Low | 11–26 | 10–26 | ✓ 96.2% agree | Various |

*Green rows = features in top-6 of BOTH methods (Jaccard=0.50). Single conflict (PreOpTLC): LIME assigns positive local contribution while SHAP assigns near-zero negative mean contribution — consistent with the methodological difference between local perturbation (LIME) and global Shapley averaging (SHAP). Spearman ρ=0.440, p=0.024; Kendall τ=0.357, p=0.011.*

### **Table S4. Gate Majority Threshold Comparison**

*Comparison of three gate majority settings on the validation cohort (n=233, 13 deaths, 220 survivors). All settings use the same model weights, gamma=0.10, and T_screen=0.6987. Majority=3/4 deployed based on optimal balance between sensitivity, alert burden, and clinical defensibility. FP/TP = false positives per true death. Alerts/100pts = (TP+FP)/233×100. Note: With EPV=0.19 (13 validation deaths, 67 features), these gate comparisons should be interpreted cautiously; differences in FP counts reflect the specific validation cohort and require external confirmation (Varoquaux 2018; Christodoulou et al. 2019).*

| **Gate** | **TP** | **FP** | **FN** | **TN** | **Sensitivity** | **Specificity** | **J** | **FP/TP** | **Alerts/100pts** | **Assessment** |
| --- | --- | --- | --- | --- | --- | --- | --- | --- | --- | --- |
| 2/4 | 13 | 56 | 0 | 164 | 100.0% | 74.5% | 0.745 | 4.3x | 29.6 per 100 | Permissive — high FP |
| **3/4 ✓** | **13** | 37 | **0** | 183 | **100.0%** | **83.2%** | **0.832** | 2.8x | 21.5 per 100 | **DEPLOYED — optimal balance** |
| 4/4 | 10 | 6 | 3 | 214 | 76.9% | 97.3% | 0.742 | **0.6x** | 6.9 per 100 | Unanimous — 3 deaths missed |

*Note: 4/4 unanimous gate is equivalent to the T_high=0.8649 HIGH_RISK zone (same 10 patients flagged). It achieves FP/TP=0.6x but misses 3 deaths (FN=3) — clinically unacceptable as a single-threshold system. The deployed 3/4 majority achieves FN=0 while maintaining acceptable alert burden (FP/TP=2.8x, 21.5 alerts per 100 patients). majority=2 flags 56 survivors (29.6 per 100) — alert fatigue risk. ✓ = deployed setting.*

References cited in supplementary: Varoquaux G. NeuroImage. 2018;166:438-443. Christodoulou E et al. J Clin Epidemiol. 2019;110:12-22. Lin TY et al. ICCV 2017:2999-3007. Gal Y, Ghahramani Z. ICML 2016;48:1050-9. Wen Y et al. ICLR 2018. Full references in main manuscript.

**END OF SUPPLEMENTARY MATERIAL**
